# Increased hazard of mortality in cases compatible with SARS-CoV-2 variant of concern 202012/1 - a matched cohort study

**DOI:** 10.1101/2021.02.09.21250937

**Authors:** Robert Challen, Ellen Brooks-Pollock, Jonathan M Read, Louise Dyson, Krasimira Tsaneva-Atanasova, Leon Danon

**Affiliations:** College of Engineering, Mathematics and Physical Sciences, University of Exeter, Exeter, Devon, UK.; Somerset NHS Foundation Trust, Taunton, Somerset, UK.; Joint Universities Pandemic and Epidemiological Research (JUNIPER) consortium.; University of Bristol, Bristol Veterinary School, Langford, Bristol, UK.; Bristol Medical School, Population Health Sciences, University of Bristol, Bristol, UK.; Lancaster Medical School, Lancaster University, Bailrigg, Lancaster, UK.; The Zeeman Institute for Systems Biology & Infectious Disease Epidemiology Research, School of Life Sciences and Mathematics Institute, University of Warwick, Coventry, UK.; The Alan Turing Institute, British Library, 96 Euston Rd, London, UK.; Department of Engineering Mathematics, University of Bristol, UK.

**Keywords:** COVID-19, SARS-CoV-2, B.1.1.7, VOC-202012/01, mortality

## Abstract

**Objectives:** To establish whether there is any change in mortality associated with infection of a new variant of SARS-CoV-2, designated a Variant of Concern in December 2020 (VOC-202012/1) compared to that associated with infection with circulating SARS-CoV-2 variants.

**Design:** Matched cohort study. Cases are matched by age, gender, ethnicity, index of multiple deprivation, lower tier local authority region, and sample date of positive specimen, and differing only by detectability of the spike protein gene using the TaqPath assay - a proxy measure of VOC-202012/1 infection.

**Setting:** United Kingdom, community - based (Pillar 2) COVID-19 testing centres using the TaqPath assay.

**Participants:** 54,906 pairs of participants testing positive for SARS-CoV-2 in Pillar 2 between 1st October 2020 and 29th January 2021, followed up until the 12th February 2021.

**Main outcome measures:** Death within 28 days of first positive SARS-CoV-2 test.

**Results:** There is a high probability that the risk of mortality is increased by infection with VOC-202012/01 (p <0.001). The mortality hazard ratio associated with infection with VOC-202012/1 compared to infection with previously circulating variants is 1.64 (95% CI 1.32 - 2.04) in patients who have tested positive for COVID-19 in the community. In this comparatively low risk group, this represents an increase from 2.5 to 4.1 deaths per 1000 detected cases.

**Conclusions:** If this finding is generalisable to other populations, VOC-202012/1 infections have the potential to cause substantial additional mortality compared to previously circulating variants. Healthcare capacity planning, national and international control policies are all impacted by this finding, with increased mortality lending weight to the argument that further coordinated and stringent measures are justified to reduce deaths from SARS-CoV-2.

## Introduction

A new lineage of the SARS-CoV-2 virus (named B.1.1.7) was identified from genomic sequencing of cases in the South East of England in early October 2020. It was identified as a variant of concern of the SARS-CoV-2 virus [1] (VOC-202012/1 - ‘new variant’) in December 2020 by Public Health England. Over December it spread from the South East to London and the rest of the UK, with three quarters of infections being attributable to the new variant by 31st December 2020 [2]. The UK implemented a second national lockdown (5/11/2020 - 2/12/2020) which coincided with the relative growth of VOC-202012/1. Following the lockdown additional control measures were implemented as the increased rate of spread of the new variant became apparent and was made public [3]. International restrictions on travel from the UK quickly followed, in particular to France and the rest of Europe late in December 2020 to curb spread of the new variant to other countries, despite evidence that it was already present outside the UK. Since then, VOC-202012/1 has been observed to be increasing in prevalence in Europe [4], and the US [5,6].

Conveniently, multiplex target Polymerase Chain Reaction (PCR) tests used in the parts of the UK national testing system are able to distinguish VOC-202012/1 from other variants. When tested using the Thermo TaqPath system it has been shown that in the UK there is a close correlation between VOC-202012/1 cases confirmed by sequencing and TaqPath PCR results where the spike protein gene PCR target has not been detected but other PCR targets (N gene and ORF1ab gene) have. These are referred to as S-gene negative, or S-gene target failure results, and have a strong association in the UK to B.1.1.7 variant infection. S-gene negative results have subsequently been used as a proxy to track the progression of this variant in the UK [7–9,2]. This association is not necessarily as strong in different countries as other variants can also cause S-gene negative results.

Sequencing of VOC-202012/1 revealed 14 genetic mutations, 8 of which occurred in parts of the genome that code for the spike protein, responsible for cell binding [10], and which influence S-gene detection. These mutations appear to have imparted a phenotypic change to the cell binding mechanism [11,7–9,2], with the potential for increased infectivity [12,13]. The impact of the change on clinical presentation, patient outcome and, most importantly mortality, remains poorly understood.

Here, using linked data from syndromic community testing and death records, we assess whether the new SARS-CoV-2 variant is associated with a different hazard of mortality compared to previously circulating variants.

## Methods

The study primarily set out to determine if there is a change in mortality in patients testing positive for SARS-CoV-2 with PCR test results compatible with VOC-202012/1. This question is difficult to answer as, during the period under study, rates of COVID-19 cases in the UK have increased dramatically, putting hospital services under strain, which in turn affects mortality [14] and potentially biases observations of mortality.

### Data availability

Data were collected by Public Health England (PHE) into a centralised database detailing the type of test performed and the results. PHE provided anonymised data to SPI-M contributors as part of the COVID-19 response under a data sharing agreement between PHE and the authors’ institutions.

### Study design

We conducted a matched cohort study. COVID-19 incidence and burden on hospitals varies in space and time and to address this as a potential source of bias, we match closely to time and geographical location of cases, and assess the variability of our estimates when relaxing the matching criteria.

### Inclusion criteria

Individuals over the age of 30 years old who had a single positive COVID-19 test result, during the period from 1st October 2020 until 29th January 2021 were included. These were restricted to test results that reported a PCR cycle threshold (CT) value. Antigen swab tests in the UK are carried out through two routes: Pillar 1 represents National Health Service (NHS) testing for those with a clinical need and healthcare workers; and Pillar 2 represents community testing of symptomatic individuals. Community based COVID-19 diagnoses are generally in a younger population with less severe disease than hospital based COVID-19 diagnoses, as elderly or severe cases tend to present directly to hospital (for more discussion see the supplementary materials). We consider only the subset of Pillar 2 tests that were processed in the high-throughput “Lighthouse” laboratories that employ the Thermo TaqPath COVID-19 multiplex Polymerase Chain Reaction (PCR) assay which amplifies the open reading frame 1a/b junction (ORF1ab) and the N and S-genes of SARS-CoV-2. We included people that had a single positive PCR test using the TaqPath assay and for which PCR cycle threshold (CT) values for the S, N and ORF1ab components of SARS-CoV-2 were available.

### Data processing

We classified the results as S+N+ORF1ab+ (“S-gene positive” - compatible with previous variants) for results that had the following CT values: S-gene < 30; N gene < 30; ORF1ab gene < 30. We classified S-N+ORF1ab+ (“S-gene negative” - compatible with VOC-202012/1) for results that had CT values: S-gene not detected; N-gene < 30; ORF1ab gene < 30. All other combinations of known CT values were classified as “Equivocal” and excluded from further analysis.

We linked the line list of case details and line list of details of death (if the patient died) using a unique study identifier. The deaths line list enumerates deaths in both hospital and community settings, and captures all deaths within 28 days of a positive COVID-19 test, following the Public Health England definition of death as ‘a death in a person with a laboratory-confirmed positive

COVID-19 test and died within (equal to or less than) 28 days of the first positive specimen date’ [15]. Maintained by Public Health England, it represents the most timely and complete record of deaths due to COVID-19 in England [15]. The deaths line list also contains some details about the timing of hospitalisation in those people that died. Cases that could not be linked and were therefore uninformative for S-gene status, were classified as “Unknown”, and were also excluded; these are generally samples not processed in Lighthouse laboratories, and include hospital cases.

During the study period hospitals experienced a period of intense demand in areas where there were large outbreaks of VOC-202012/01, which potentially adversely impacted patient outcomes. To control for any systematic bias this could have introduced, we paired individuals with S-gene positive test results (“non-exposed”) to individuals with S-gene negative test results (“exposed” - highly likely to be VOC-202012/01) by matching on gender, ethnicity, index of multiple deprivation (IMD), location (as lower tier local authority region ~ 190,000 people), age (within a tolerance of ±5 years), and date of positive specimen (within a tolerance of ±1 day).

Some S-gene negative cases matched multiple S-gene positive cases and vice versa, so we sampled them randomly within our framework to generate 50 replicates, ensuring no S-gene negative or S-gene positive was present more than once in each replicate. All analyses were conducted on each replicate as a separate sample and the results combined by combining the beta-coefficient estimates as a mixture of normal distributions, and calculating combination mean and confidence intervals numerically from the mixture distribution (further discussion of this topic is found in the supplementary materials).

### Statistical analysis

Cases were followed up for 28 days following infection or until 12 February 2020, after which point cases that had no record of death were censored.In these data over 50% of COVID-19 related deaths are reported within 3 days of date of death and over 95% within 14 days [16] (more details available in the supplementary materials). There are no differences in the reporting patterns for S-gene negatives, and S-gene positives. The deaths line list is constructed from multiple sources as the gold standard list of COVID-19 related mortality in England, and ultimately will include all deaths where COVID-19 is mentioned on the death certificate. We compared the rates of death in our community-based dataset between S-gene positives versus S-gene negatives. We calculated the hazard ratio of death given a S-gene negative test result, versus death given a S-gene positive test result using a Cox proportional hazards model [17] with age (years) as a linear covariate, taking into account censoring. All analyses were performed in R (version 3.6.3)[18–20] and all analysis code is available (doi:10.5281/zenodo.4543510).

### Sensitivity analyses

We examined different inclusion criteria for sources of systematic bias. We systematically adjusted values for cycle thresholds for the S, N and ORF1ab genes, and the tolerances of our algorithm to match both inexact age and inexact specimen dates.

## Results

There were 941,518 patients older than 30 with a single positive TaqPath test between 1st Oct 2020 and 28th January 2021. From these we identified 214,082 cases who match at least one other case with similar ages and specimen dates, and identical gender, ethnicity, geography and index of multiple deprivation, and differing only by S-gene status. Sampling these pairs to ensure they represented unique patients resulted in 50 replicates with an average of 54,906 S-gene positive cases and 54,906 S-gene negative cases per replicate. Every case was followed up for a minimum of 14 days after the positive test result, and over 85% of the cases were followed for the whole 28 day period (more details available in the supplementary materials). Of these average 109,812 patients, 367 died (averaged over the 50 replicates) within 28 days of a positive test (0.24%) - see Table 1. The matching and sampling process is observed to control well for all demographic and geographic variables considered (with slight mismatches due to differences in scale from matching and reporting). With age and specimen date, where we allowed small tolerances, the average difference between ages in the S-gene positive and S-gene negative arms was 0.0 years and a mean difference of 0.2 days was observed between specimen dates (with S negative specimens taken later than S positives).

**Table 1:**
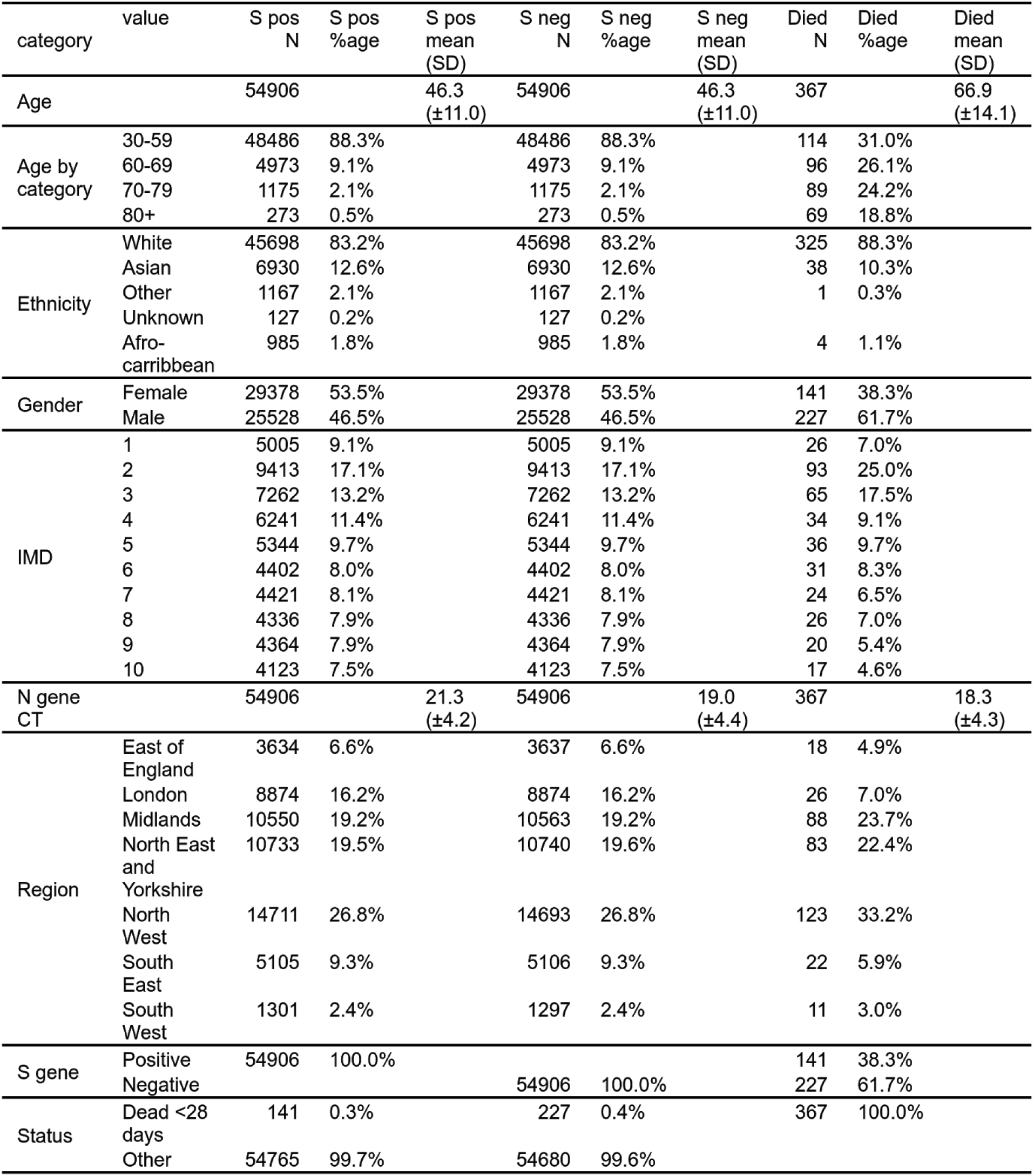
S-gene positive (“non-exposed”) & S-gene negative (“exposed”) matched cases based on age, ethnicity, gender, index of multiple deprivation, geography and specimen date (not shown). For comparison the subset of exposed and non-exposed who died are presented in the right 3 columns. IMD is an index of multiple deprivation.

We observe that the subset of cases who go on to die are generally older (mean 66.9 years old in deaths versus 46.3 years old in all cases), and a higher proportion are men, as has been reported by previous analysis [21]. We note both cases and deaths are under-represented in the South West and East of England, since these areas have not used TaqPath assays until recently, and thus do not report S-gene status.

We found an average of 227 deaths out of 54,906 patients in the S-gene negative “exposed” arm of the study compared to 141 out of 54,906 in the S-gene positive “non-exposed” arm. This gives a hazard ratio of 1.64 (95% confidence intervals 1.32 - 2.04; p = <0.001) over the whole follow up period - see table 2. The rate of death of S-gene negatives and S-gene positives over time diverges after 14 days, shown in figure 2. This is graphical evidence that the hazard ratio is not constant over time, and as such this violates the proportional hazards assumption of the Cox model. This was investigated further (details presented in the supplementary materials) and this violation may be corrected by considering the hazard ratio in days 0 to 14 of follow up, versus that in days 15 to 28. The hazard ratio in the first period was not significantly elevated, but in days 15 to 28 the hazard ratio is found to be 2.40 (95% CI 1.66-3.47).

**Table 2:** Hazard ratios for death given an S-gene negative test result versus deaths given a S-gene positive result (reference category). Hazard ratios greater than one are indicative of an increased rate of death in infections compatible with VOC-202012/01. In the first model we look at the S-gene status as an indicator with age as a covariate, in the second model we include variability in the N gene CT value measured in the original specimen as a continuous predictor, which explains some but not all of the hazard ratio increase observed due to S-gene negativity.

| Model | Predictor | Value | Hazard ratio (95% CI) | p value |
| --- | --- | --- | --- | --- |
| S gene + age | S gene status | Positive (ref) | — | — |
|  |  | Negative | 1.64 (1.32 – 2.04) | <0.001 |
|  | Age (per decade) |  | 3.55 (3.28 – 3.84) | <0.001 |
| S gene + N gene CT + age | S gene status | Positive (ref) | — | — |
|  |  | Negative | 1.37 (1.09 – 1.72) | 0.004 |
|  | Age (per decade) |  | 3.51 (3.24 – 3.80) | <0.001 |
|  | N gene CT (per 10 units) |  | 0.50 (0.39 – 0.65) | <0.001 |

**Figure 1.**
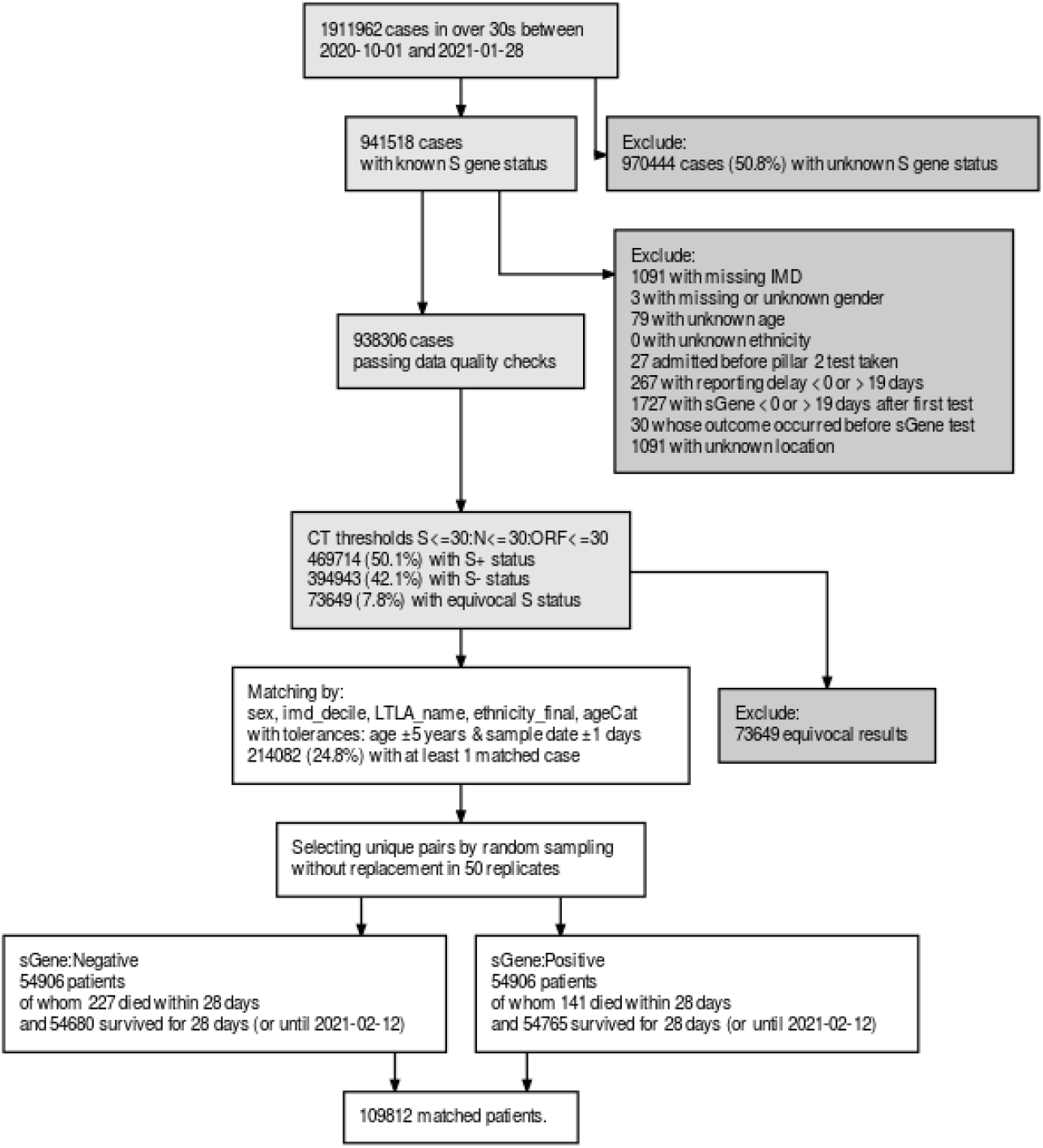
the sample selection algorithm. In the matching process we randomly sample to create 50 replicates. In this figure we have given average figures for the numbers of patients in each arm of the study. LTLA is a geographical location as a lower tier local authority, IMD is index of multiple deprivation.

**Figure 2.**
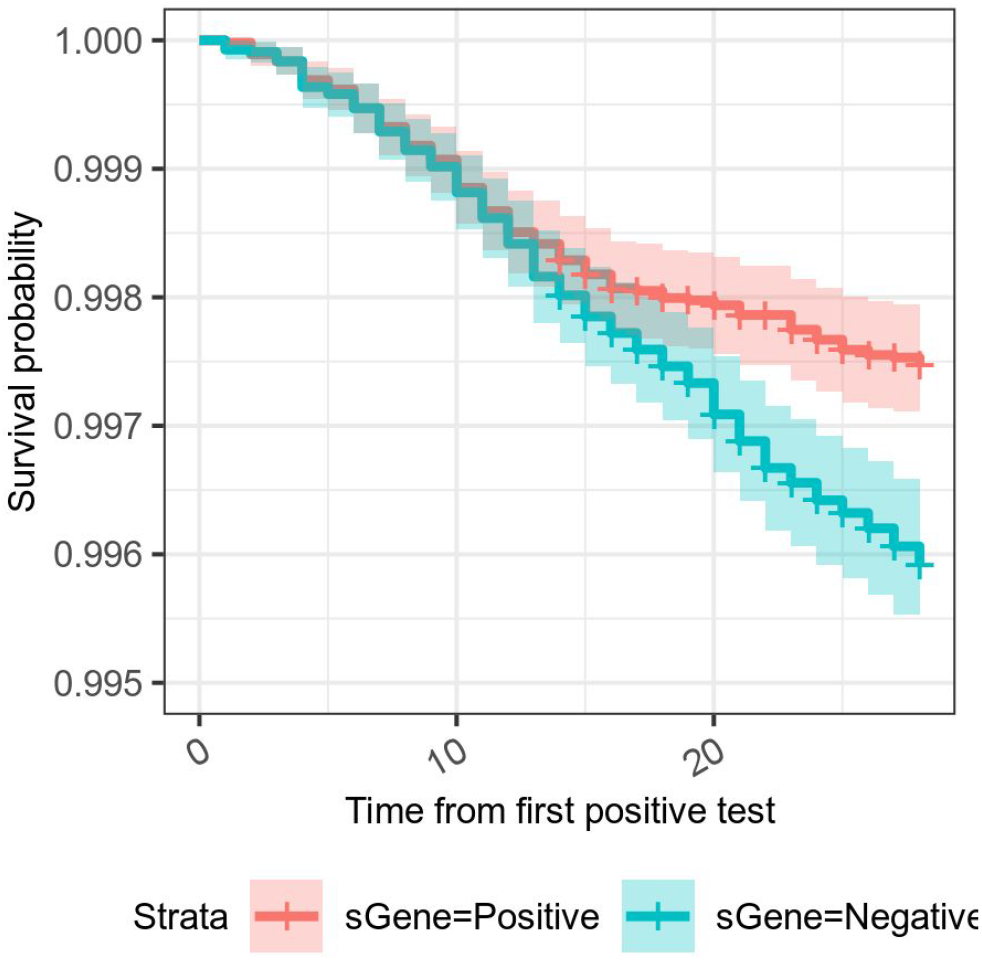
A Kaplan-Meier survival curve for S-gene positive (previously-circulating variants) and S-gene negative (VOC-202012/1) infections in the UK. The y axis has been clipped as mortality in both groups is low.

The matched cohort design controls for most potential biases including variations in hospital capacity, as it pairs patients by demographics, geography and time of testing. We investigated other further potential biases that may be present. One possibility for bias could be a difference in the timing of the presentation of S-gene negative and S-gene positive infections for testing, with for example S-gene positives presenting earlier, and thus appearing to progress slower. We only have hospital admission data for patients who ultimately died but in this there is no evidence for asymmetric delays in time from test to hospital admission as shown in figure 3 panel A. This has also been investigated by the Office of National Statistics who find that S-gene negative patients are more likely to present earlier for testing [22].

**Figure 3.**
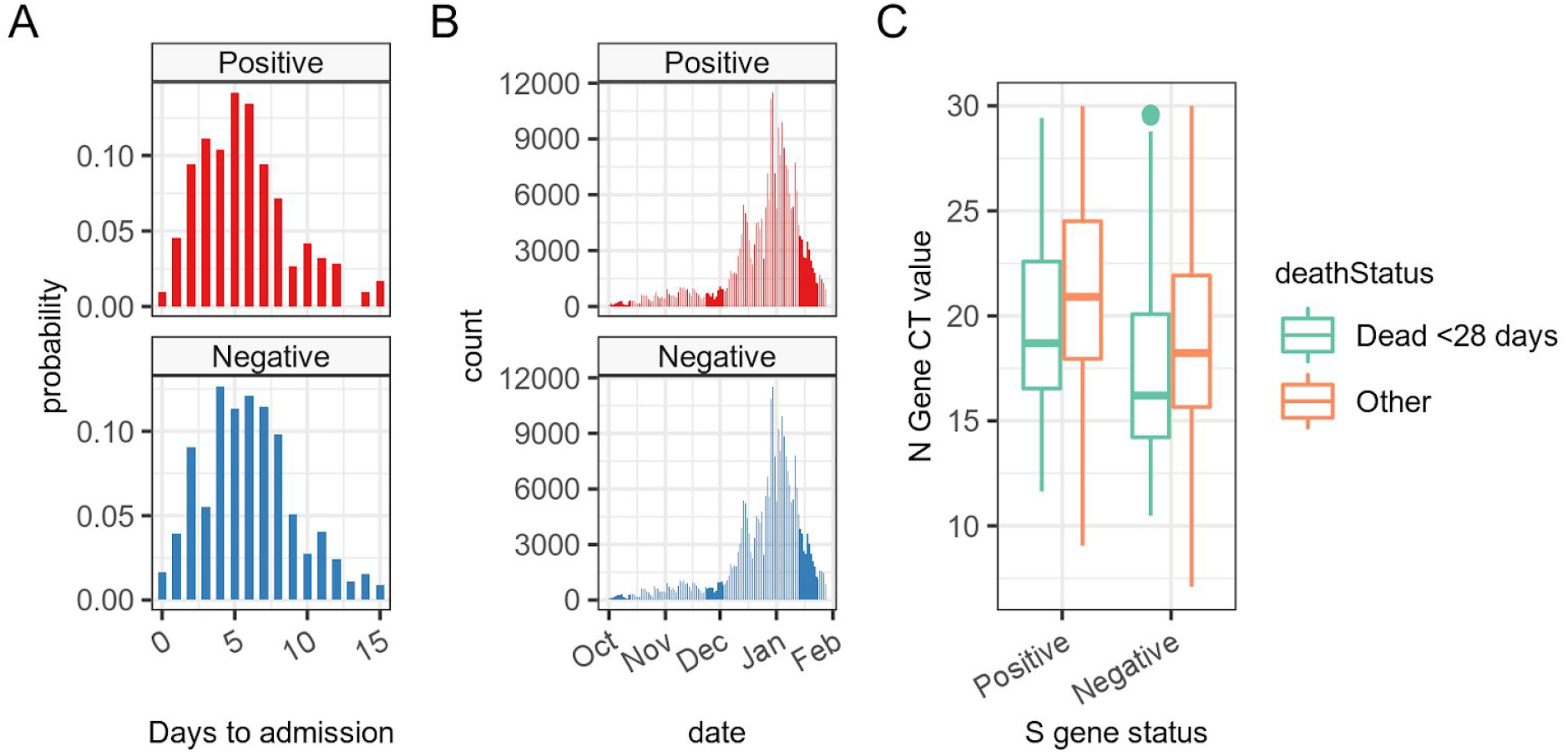
Investigation of biases in the exposed and non-exposed arms. In panel A we see no discernible pattern in the delay between specimen date and hospital admission in patients who subsequently died. In panel B we see the date distribution of the specimens in our matched pairs of S-gene positives and negatives - concentrated in December and January. In panel C we see median N-gene CT values for both S-gene positives and negatives, demonstrating lower CT values in the initial tests of those who subsequently died, and in those with S-gene negative infections.

As the degree of hospital load in the UK varied considerably over the study period it is important to note that there is no avoidable temporal clustering in the matched cases. Figure 3 panel B demonstrates that the paired cases we investigated are spread throughout time, but concentrate around the end of December 2020 and beginning of January 2021. As the ratio of S-gene negative and S-gene positive cases changes over this period, in the early stages it is hard to find S-gene negatives for S-gene positive equivalents, and in the later stages hard to find S-gene positives for S-gene negative equivalents, resulting in the bulk of our matched cases being during the time of transition from S-gene positive variant dominance to S-gene negative variant dominance (discussed further in the supplementary materials).

We note in table 1 and in figure 3 panel C that CT values for the N-gene are lower in S-gene negatives than in S-gene positives, and this effect is potentiated in those who died. Low values for the N-gene cycle threshold imply the viral load in patients at the time of sampling was higher. The higher viral load of S-gene negatives could be the biological result of the VOC202012/1 mutations and be a cause of higher mortality, but alternatively it could be an indication of the timing of testing, with S-gene negatives presenting at peak infectiousness, for some as yet unknown reason. Thus N-gene CT values could be regarded either as an indication of bias or as a feature of S-gene negative infection. If we interpret it as a source of bias, we can control for N-gene CT value in the Cox proportional hazards model (in table 2 - second model) which shows a reduction in the overall hazard ratio due to S-gene negativity to 1.37 (95% CI 1.09 - 1.72). Even if we do not consider elevated viral load as a biological feature of S-gene negative infection, the residual increase in hazard ratio implies a mortality effect not explained by viral load alone.

### Sensitivity analysis

Figure 4 shows the estimates of hazard ratios related to the alteration of assumptions taken in our analysis. In panel A we see that reducing the CT value threshold for identifying a particular gene leads to a reduction in the central estimate of hazard ratios. Lower CT thresholds reduce the number of certain S-gene positive and S-gene negative results and increase the number of equivocal results, which are subsequently excluded from the analysis, leading to an effective reduction in overall case numbers. Given that CT values are generally higher in true S-gene positive cases, reducing the CT threshold will tend to reclassify more mild S-gene positive cases than mild S-gene negative cases as equivocal. This could explain the small reduction in hazard ratio associated with reducing the CT threshold. There appears to be a marginal, non-significant rise at a CT threshold of 30; we choose this as our central estimate since this is a standard CT threshold employed by most laboratories.

**Figure 4.**
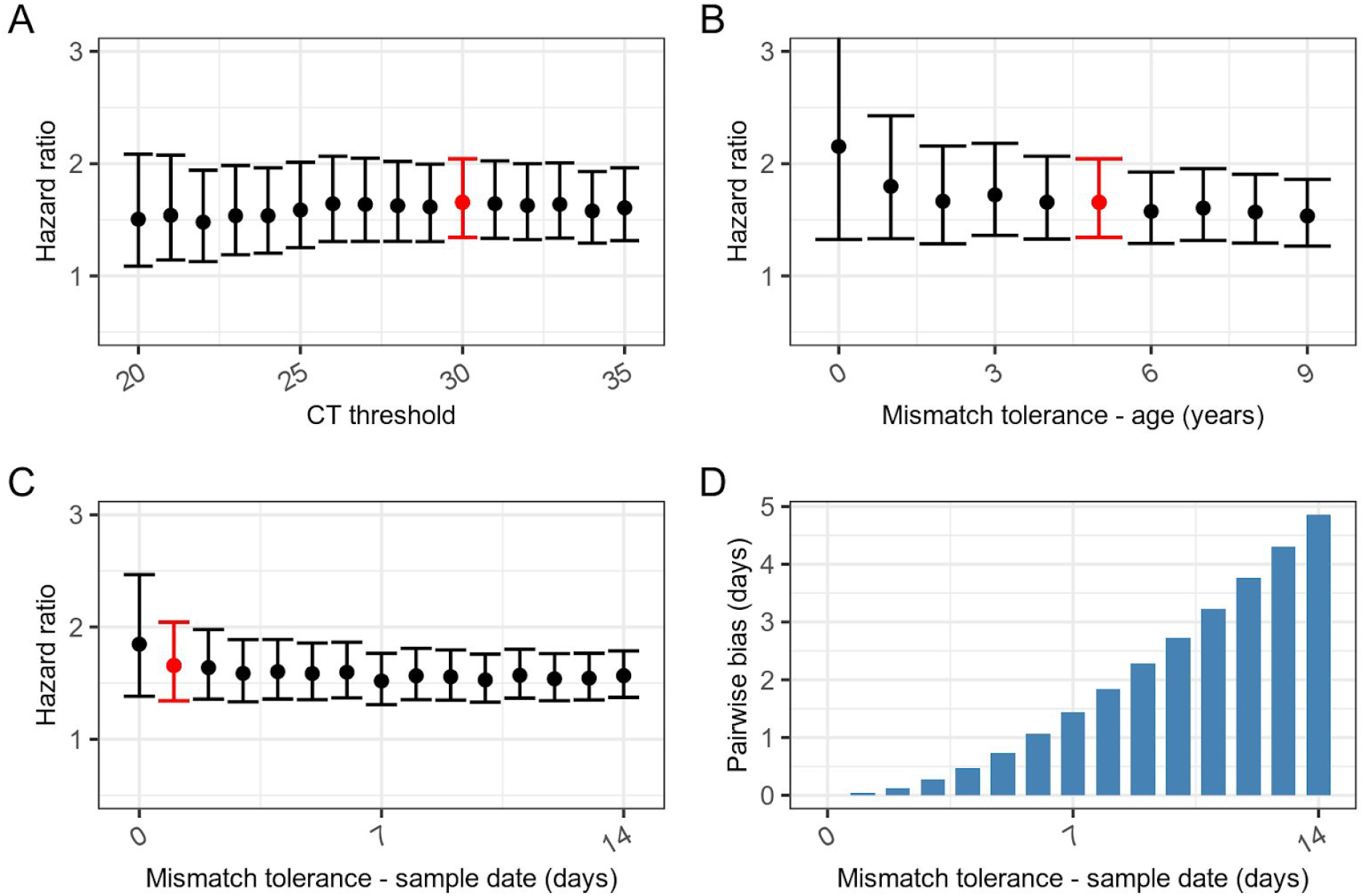
Sensitivity analysis. Panel A - central HR estimates are mildly affected by the threshold for CT values used in definition of gene positivity. Panel B - allowing greater flexibility in age when selecting cases leads to small changes in the associated HR estimate, and is included as a covariate in the Cox model. Panel C - allowing greater flexibility in sample date when selecting paired cases leads to slightly lower estimates of hazard ratios, however because the prevalence of exposed and non-exposed changes over time, it also results in increasing systematic bias between pairs of exposed and non-exposed (labelled “pairwise bias” in panel D), where in the most permissive matching S-gene negatives could be on average up to 5 days later that S-gene positives. In panels A,B & C the red bar indicates our default assumptions (CT < 30; age tolerance ±5 years; sample date tolerance ±1 day) from the rest of the paper.

Panel B shows the effect of allowing age to be mismatched between exposed and non-exposed. This does not have the same systematic bias as sample date and the mean age difference between exposed and non-exposed is less than 0.005 years. Age is a strong predictor of mortality in COVD-19, so we might expect some potential bias, however we control for this when calculating the hazard ratios (see Table 2).

Panel C shows the effect of allowing greater time between sample dates in matched S-gene positives and S-gene negatives, increasing the temporal uncertainty and diluting the hazard ratio. Because of the change in prevalence from predominantly S-gene positive to predominantly

S-gene negative, increasing the tolerance to sample date mismatch leads to a systematic “pairwise bias” in the dates of the original positive test result (Panel D) with S-gene negatives generally being identified after S-gene positives. Given that over the study period cases were exponentially increasing this risks introducing a bias if hospital capacity begins to be exceeded over time. For this reason, we aimed to minimise the sample date tolerance, trading off the reduction in bias against the variance introduced by the reduced case numbers resulting from tight matching criteria.

Despite the differences between all the combinations investigated, all studies report a statistically significant rise in the mortality hazard ratio imparted by the variant, suggesting a real effect, and the vast majority of central estimates are within the range of 1.5 to 1.7. Please see the supplementary materials for further discussion of other potential covariates.

## Discussion

VOC-202012/1 infections (as measured by S-gene negativity) are associated with an elevated risk of death (p<0.001) in people testing positive for COVID-19 in the community. The increased hazard ratio (HR) between 1.32 and 2.04 over and above other variants, translates to a 32% to 104% increase in mortality, with the most probable HR estimate of 1.64, or a 64% increase. However, the absolute risk of death in this group of community-identified cases remains low, rising from 2.5 to 4.1 deaths per 1000 cases.

The matched cohort approach controls for many of the biases we are aware of. In particular, mortality is affected by how many cases require intensive care in a hospital setting [14]; increasing numbers of cases in the study period (1st October 2020 - 12th February 2021), compounded by staff absenteeism due to COVID-19 infection or isolation due to contact with infected individuals, has placed intense strain on hospital services and a reduction in the staff to patient ratio. This may have affected mortality and is a potential source of bias. This is controlled for by matching individuals by administrative region and time of positive test (within 1 day), which constrains pairs to receive care at the same place and time, and we assert a similar level of care. Age-related mortality is controlled for by matching by age (within 5 years), but is also controlled for by the proportional hazards model employed.

This is a community-based study. We do not have information about the S-gene status of patients in hospitals. The community-based testing (Pillar 2) in this dataset covers a younger age group and hence represents less severe disease than cases detected through hospital-based testing (Pillar 1). In cases detected in the community, death remains a comparatively rare outcome, compared to in-hospital identified cases. Our study only includes approximately 8% of the total deaths occurring during the study period. Of all coronavirus deaths, approximately 26% occur in individuals who are identified in the community, and only 30% of these have S-gene data [23]. Whether the increase in mortality in community cases is also observed in elderly patients, or hospitalised cases, remains to be seen.

We cannot exclude a selection bias. Community testing is largely self-selected, or driven by contact tracing. There remains a potential bias if there were a higher proportion of undetected asymptomatic cases in S-gene negative infections than in S-gene positive infections. In this event, VOC-202012/1 cases may be at a more advanced stage of infection when detected, and have a higher apparent mortality. This could be consistent with the lower N-gene CT values observed in S-gene negatives. Our analysis, or any retrospective study based on symptomatic cases, would not be able to detect this, however early survey data suggests that S-gene negative infections are, if anything, more likely to present for testing [22]. Addressing this potential bias requires a study design capable of detecting asymptomatic infections in S-gene positives and S-gene negatives.

Some of the increased hazard could be explained by comorbidities. There is no information about comorbid conditions in the data we analysed, although this will be partly controlled by age, ethnicity and index of multiple deprivation. There is currently no evidence of a mechanistic reason why people with certain comorbidities would be infected with one variant or the other. However, it is possible that people with certain comorbidities are both at a higher risk of infection with VOC-202012/1 and have a higher mortality rate. This would tend to reduce the hazard ratio attributable to VOC-202012/1 alone.

Our preliminary estimate of the hazard ratio was 1.91 (95% CI 1.35 - 2.71), which is marginally higher than the estimate presented here with compatible uncertainty [23,24]. This was based on 94 S-gene negative deaths and 49 S-gene positive deaths in 66,208 less strictly matched pairs. As the situation has unfolded and more data has become available we have been able to narrow our tolerance for mismatches, and increase the proportion of cases with complete follow up, to get more accurate central estimates. The design of this study is well suited to determining whether there is an elevated risk of death, in an unbiased manner, but uses a comparatively small number of patients. Other study designs, involving the use of wider unpaired samples, may be better at quantifying the absolute increase in risk, but with more potential for bias [25]. Other recent studies produced similar estimates of the increased hazard ratio. These studies use the same community-based testing data but have employed different study and analysis designs. Their preliminary results were compatible point estimates of the mortality hazard ratio (1.3 - 1.65), and the confidence intervals of these studies overlap with those described here [23]. As with our work, these other estimates are being continuously re-evaluated as more data is acquired and in subsequent updates some of these have been revised upwards [26].

## Conclusion

The variant of concern, in addition to being more transmissible, appears to be more deadly. We expect this to be due to changes in its phenotypic properties associated with multiple genetic mutations [27], and see no reason why this finding would be specific to the UK. This concerning development, borne out in epidemiological analyses, implies that there will be an increase in the rate of serious cases requiring hospital attention. At the time of writing (15th February 2021) the national lockdown appears to be effective at reducing the transmission rate in the UK, but control of the outbreak has been made more difficult by the proliferation of the new variant. The resulting number of deaths will scale linearly with the proportion of cases infected with the new variant. Other analyses have indicated that the new variant is also associated with increased transmissibility, which would lead to a potentially exponential increase in the resulting number of deaths [12]. Clinicians at the front line should be aware that a higher mortality rate is likely even if quality of practice remains unchanged. This has broader implications for any vaccination allocation policy designed to reduce mortality in the late-middle age-groups typical of the community-identified cases in this dataset.

The question remains whether excess mortality due to VOC-202012/1 will be observed in other population groups, particularly the elderly, care home residents, and those with other comorbidities who generally present directly to hospitals as emergencies. Hospital-based studies require a mechanism to distinguish emerging variants from previously circulating variants, currently only done through genotyping. Due to the effort involved, the proportion of genotyped samples representing hospitalised cases remains low, and we argue that PCR tests that specifically target VOC-202012/1 mutations should be more widely used.

Moreover, the emergence of VOC-202012/1 and its mutations (including E484K), combined with other variants of concern including those identified in Brazil and South Africa [28] highlights the capacity of SARS-CoV-2 to rapidly evolve new phenotypic variants, with vaccine escape mutants being a real possibility [29]. Our method helps characterise the clinical presentation and outcome of one new variant, but is generalisable to others, given sufficient amounts of informative data.

However, assessment of the clinical outcomes of multiple circulating phenotypic variants requires scalable technology capable of identifying substantial numbers of cases due to emerging variants (e.g. broad PCR assay panels targeting variant foci [30]) and robust collection of outcome data.

The effect of time, location, case rates, age and treatment pathways have been controlled for in this study, but are important factors to understand if we are to improve future outcomes. Future work on the relative impact of these may allow for better targeting of resource allocation [31], vaccine distribution strategies and, in the future, relaxation of restrictions.

## Data Availability

The data are available on request and appropriate data sharing agreement from Public Health England. Code for the analysis is available at doi: 10.5281/zenodo.4543510

https://doi.org/10.5281/zenodo.4543510

## Authors contributions

RC and LDanon designed the study, all authors discussed the concept of the article and RC performed analyses and wrote the initial draft. KTA, EB-P, LDanon, JMR and LDyson commented and made revisions. All authors read and approved the final manuscript. RC is the guarantor. The views presented here are those of the authors and should not be attributed to Somerset Foundation NHS Trust or the GDE.

## Acknowledgements

We extend our thanks to Professor David Spiegelhalter for comments on an early draft, and to Dr Nick Gent at Public Health England for providing access to the data.

Patient and public involvement: Due to the nature of this research there was no involvement of patients or members of the public in the design or reporting of this study. Direct dissemination to study participants is not possible.

## Data sharing

Data are available to members of universities that have data sharing agreements in place with Public Health England.

## Ethics

The data were supplied from the SGSS database and death reports after anonymisation under strict data protection protocols agreed between the University of Exeter and Public Health England. The ethics of the use of these data for these purposes was agreed by Public Health England with the Governments SPI-M(O) / SAGE committees. The research was assessed as not needing NHS Research Ethics Committee review.

