## Supplementary materials for "Increased hazard of mortality in cases compatible with SARS-CoV-2 variant of concern 202012/1 - a matched cohort study"

**Assessment of potential data biases** 1

**Case Matching** 3

**Combining estimates** 4

**Additional Proportional Hazards Models** 5

**Proportional hazards assumption** 7

**Supplementary material references** 8

#### Assessment of potential data biases

In our analysis the percentage of COVID-19 cases that die within 28 days of a positive test appears lower than would be expected from a simple Infection Fatality Ratio (IFR) calculation of all COVID-19 cases. One reason for this is that the community-testing dataset we are using (Pillar 2) does not include testing in healthcare settings, and so represents a younger than average population. Figure S1 shows the distribution of S-gene positive, S-gene negative and equivocal S-gene cases in Pillar 2 by single year of age, compared to the distribution of people in the general population (grey). When we consider that elderly patients are less likely to be tested in the community, prior to admission to hospital, we can expect that mortality in the younger community cohort will be lower.

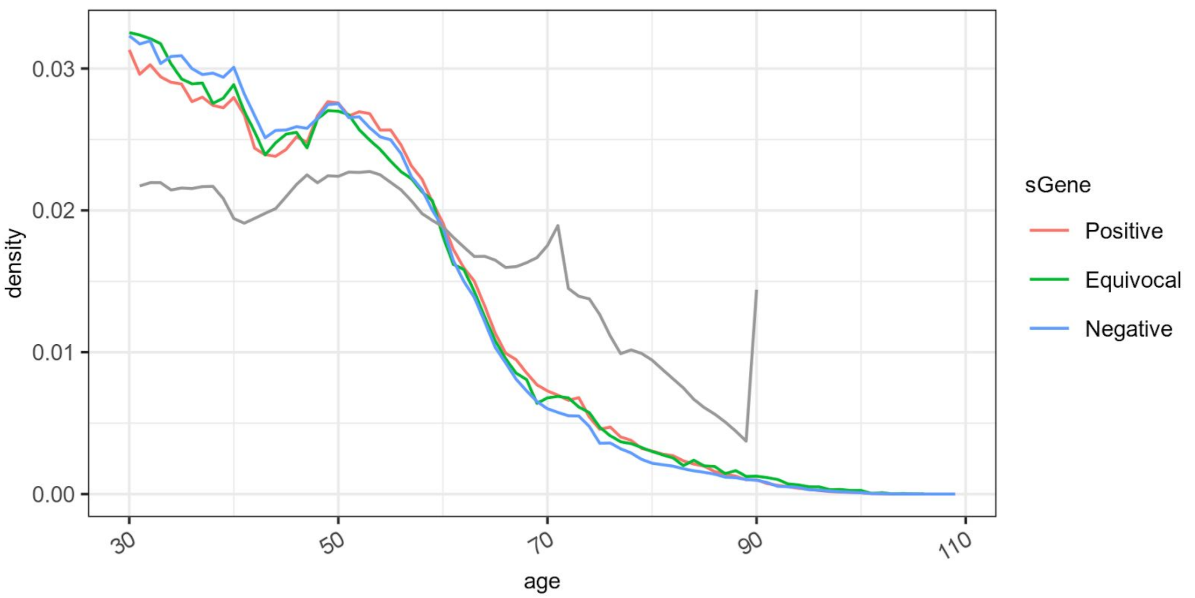


*Figure S1 - Distribution of ages for S-gene positive (red), negative (blue) and equivocal (green) cases from Pillar 2 (community testing) data, compared to the general population (grey). In this figure we take unmatched COVID-19 cases for which S-gene data is available (941,518 cases) and compare the age distribution to that in the ONS 2019 mid year estimates for the general population (which has an upper age category of 90+, and now lags the testing data by 3 years).*

A potential source of bias which could influence the estimation of the hazard ratio would occur if there was either a differential loss or a differential delay to reporting of outcome in either of the 2 arms of our matched cohort. In Figure S2 we investigate the delay in reporting of death and find the majority of all deaths are reported within 14 days, with minimal differences between S-gene positive and S-gene negative, and conclude that this source of bias is negligible.

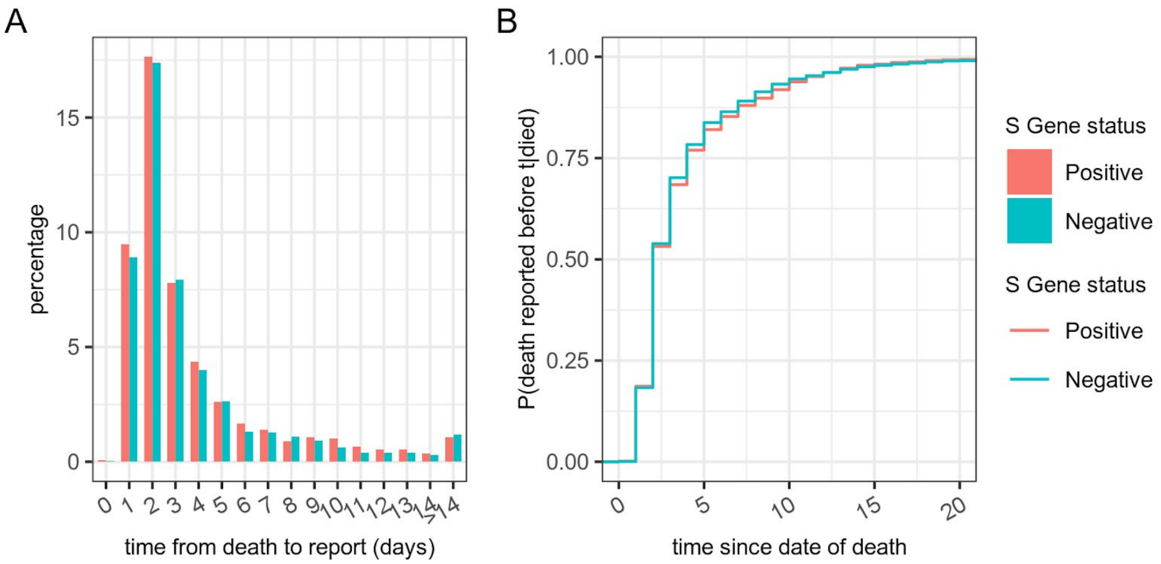


*Figure S2 - Delays from date of death and reporting of death in the dataset studied. Panel A shows the distribution of times from date of death to report of death, with the cumulative distribution shown in panel B. We see no discernable difference between the distribution for S-gene positive (red) and S-gene negative (blue) individuals. Over 50% of deaths are reported within 3 days of the date of death.*

We analyse data involving samples taken up to the 29th January 2021 and follow cases up for 28 days or the 12th February 2021 (whichever is earlier). This means that all cases have at least 14 days follow up and 85% of cases have the full 28 days as seen in Figure S3

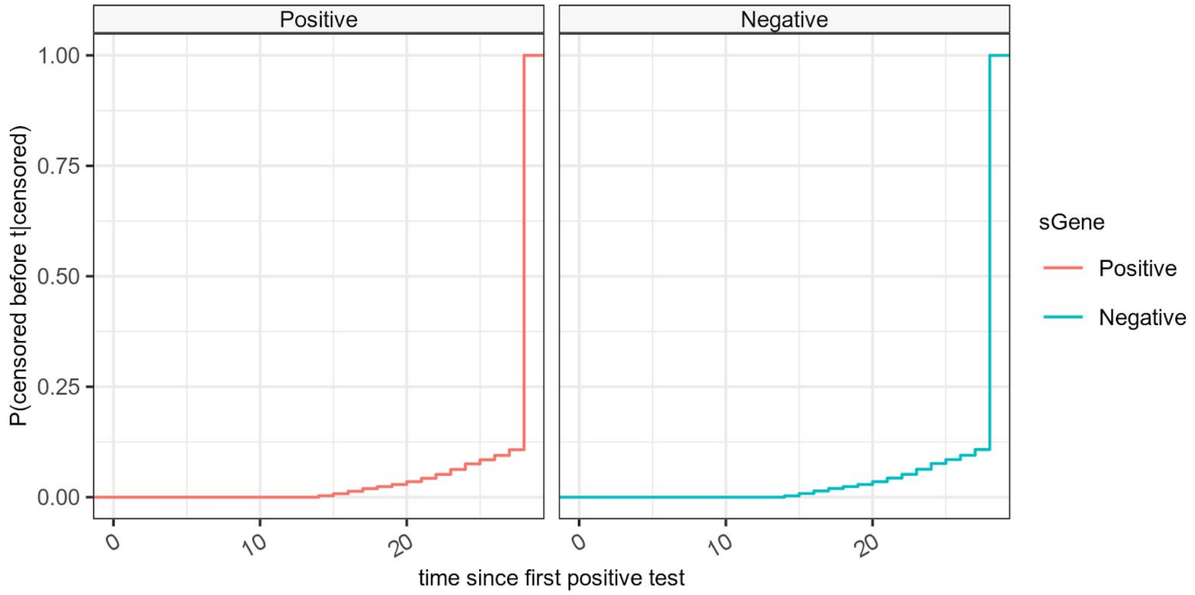


*Figure S3 - Timing of censored cases shows the cumulative proportion of cases that are censored by time since their first positive test. Less than 15% of cases are censored in this analysis and no cases are censored before 14 days. The censoring of cases is indistinguishable between S-gene negative (blue) and positive (red) cases as we are matching cases nearly exactly by sample date.*

### Case Matching

Matching of S-gene negative and S-gene positive cases in this study is an unusual problem, as unlike most other case control studies the prevalence of both S-gene negative and S-gene positive cases was dynamic over the study period. This is in contrast to a rare effects matched cohort study where rare “exposed” cases can be matched to a number of “un-exposed” from the very large general population. In our case, as the prevalence of both S-gene negative and S-gene positives varies, the ease of pairing them also varies. This results in a potential asymmetry in the pairs matched which is time dependent as demonstrated in Figure S4.

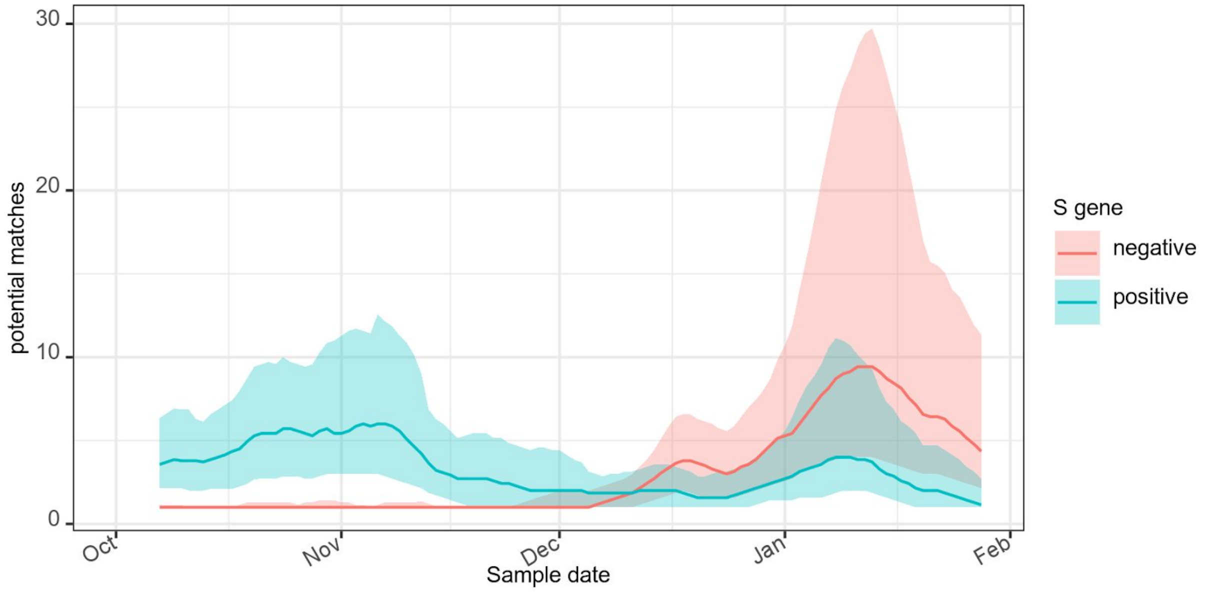


*Figure S4 - The average number and interquartile ranges of the number of potential matches for individual S-gene negative (red) or S-gene positive (blue) cases as a function of time over the study period. Initially it was much easier to find S-gene positive matches for S-gene negatives which were relatively scarce. By the end of the time series the opposite is true.*

Since individual S-gene negative cases can match multiple S-gene positive cases and vice versa it is necessary to have a strategy for selecting individuals or individual pairs from the range of possible options. We took three approaches to this. Firstly an unpaired selection strategy in which we take unique individuals out of the matched pairs into a single replicate. Secondly an edge sampling strategy in which we randomly sample pairs with replacement into multiple replicates, ensuring unique cases in each replicate, and thirdly a node sampling strategy in which we randomly sample first cases then associated paired matches into multiple replicates and ensure unique cases in each replicate.

The comparison of the unpaired cases, node and edge sampling strategies on the outcome of the hazard rate estimation is shown in Table S1. All other parameters in this analysis are the same as the central estimates in Table 2 of the main paper, which uses the node sampling strategy. All three matching strategies give results with similar estimates of hazard ratios. All models show a significantly elevated HR for S-gene negativity, and the confidence limits are very similar. The central estimates of the hazard ratio vary with different selection strategies but this variability is small compared to the confidence intervals.

**Table S1. Comparison of three methods for resolving multiple matches in the data set, on the final hazard ratio estimates. The unpaired strategy selects all unique cases that match. The edge sampling strategy selects random replicates without replacement based on pairs, and the node sampling strategy selects random replicates without replacement based on the individuals.**

| Strategy | Predictor | Value | Hazard ratio (95% CI) | p value |
|---|---|---|---|---|
| Edge sampling | S-gene status | Positive (ref) | — | — |
| | | Negative | 1.60 (1.26 – 2.03) | <0.001 |
| | Age (per decade) | | 3.54 (3.25 – 3.86) | <0.001 |
| Node sampling | S-gene status | Positive (ref) | — | — |
| | | Negative | 1.64 (1.32 – 2.04) | <0.001 |
| | Age (per decade) | | 3.55 (3.28 – 3.84) | <0.001 |
| Unpaired cases | S-gene status | Positive (ref) | — | — |
| | | Negative | 1.65 (1.37 – 1.98) | <0.001 |
| | Age (per decade) | | 3.49 (3.29 – 3.71) | <0.001 |

#### Combining estimates

Using random sampling generates 50 replicates of different case compositions, and for each replicate we fitted a set of Cox proportional hazards models, considering each replicate as an individual sample. This produces 50 differing estimates and confidence intervals for the hazard ratios and beta-coefficients of each component of the resulting mortality model. To produce a single combined estimate for the hazard of any given covariate, we assumed the estimates of beta-coefficients from the cox-models to be normally distributed and combined their probability density functions from each bootstrap replicate into a mixture distribution, as shown in red in figure S6. This mixture and its associated cumulative density function were used to determine the 95% confidence intervals, with a Newton-Raphson numerical approach (red dashed lines in figure S6).

Figure S6 shows the combined estimate of the hazard ratio of S-gene negative infection in the main model presented in our paper, and gives us a sense of the stability of our estimate in the face of the random variation introduced by the node sampling pair selection strategy. It shows our central estimate of the hazard ratio of 1.64 is the middle value of a family of estimates ranging from 1.5 to 1.8.

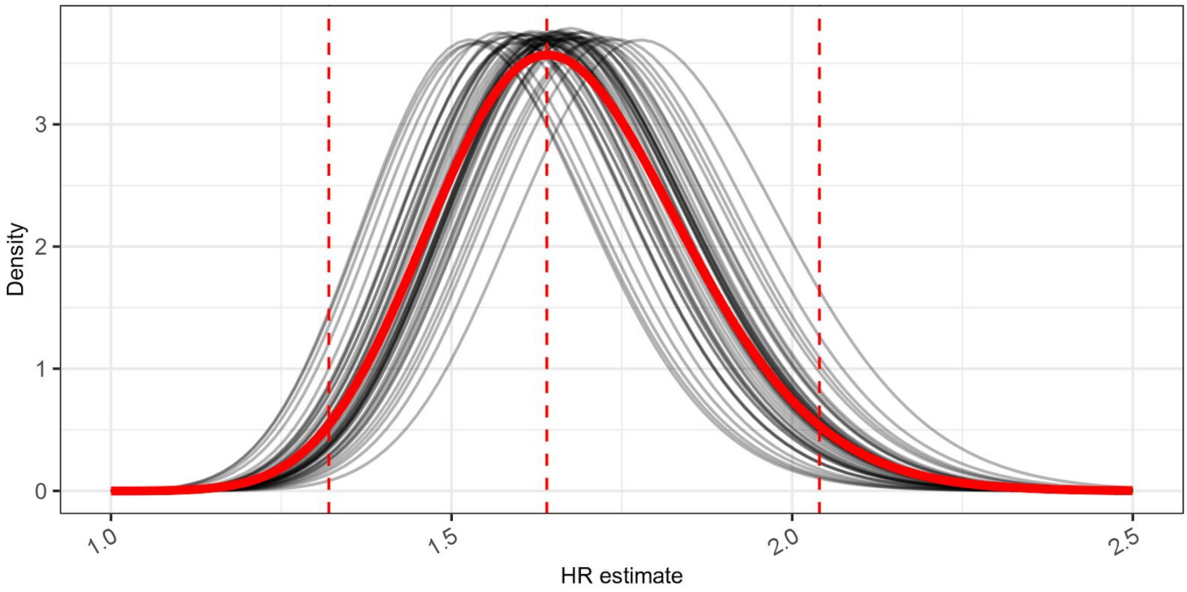


*Figure S6 - Combining point estimates from replicates was performed assuming the beta-coefficient of the cox model to be normally distributed, and summing the probability density functions to make a mixture distribution. In this figure the exponential of the beta-coefficients of the component and mixture distributions are shown as hazard ratio estimate distributions.*

#### Additional Proportional Hazards Models

To further assess the robustness of our findings, we fitted a model which included additional individual-level covariates: sex, ethnicity, and Index of Multiple Deprivation (IMD) of patient home location, on top of the S-gene status and age. We found the hazard ratio associated with negative S-gene status to be comparable to the model without the additional covariates as shown in Table S1. This is to be expected as the matched cohort approach ensures that these covariates are independently associated with mortality compared to the S-gene status. As may be expected from other studies of COVID-19 mortality, we find increasing age and male sex are signficantly associated with a higher hazard of mortality. Because we exclude many cases in the pairing process we do not have sufficient power to determine any other associations.

**Table S2. Cox proportional hazard model with additional covariates.**

| Model | Predictor | Value | Hazard ratio (95% CI) | p value |
|---|---|---|---|---|
| S-gene + covariates | S-gene status | Positive (ref) | — | — |
| | | Negative | 1.64 (1.31 – 2.04) | <0.001 |
| | Age (per decade) | | 3.62 (3.33 – 3.94) | <0.001 |
| | Ethnicity | White | — | — |
| | | Afro-caribbean | 1.03 (0.27 – 3.18) | 0.455 |
| | | Asian | 1.03 (0.71 – 1.49) | 0.427 |
| | | Other | 0.00 (1.00 – Inf) | 0.301 |
| | | Unknown | 0.00 (1.00 – Inf) | 0.496 |
| | IMD | 1 | 1.53 (0.96 – 2.46) | 0.037 |
| | | 2 | 1.68 (1.04 – 2.73) | 0.016 |
| | | 3 | 1.16 (0.68 – 2.01) | 0.292 |
| | | 4 | 1.45 (0.85 – 2.48) | 0.085 |
| | | 5 (ref) | — | — |
| | | 6 | 1.38 (0.80 – 2.39) | 0.125 |
| | | 7 | 0.92 (0.51 – 1.66) | 0.394 |
| | | 8 | 0.96 (0.54 – 1.71) | 0.440 |
| | | 9 | 0.69 (0.37 – 1.29) | 0.122 |
| | | 10 | 0.67 (0.35 – 1.27) | 0.110 |
| | Gender | Female (ref) | — | — |
| | | Male | 2.13 (1.71 – 2.67) | <0.001 |

The relationship between age and hazard ratio is most simply described by a linear term in the proportional hazards model, however it is possible that this relationship is non-linear. To test this we constructed a model which involved S-gene status and age and a restricted polynomial spline, and visualised the resulting coefficient. We conducted this analysis on a single replicate, and on the combination of all replicates. Both showed the same pattern as in Figure S7 in which it is clear that any non-linearity in the contribution of age effects is very minor.

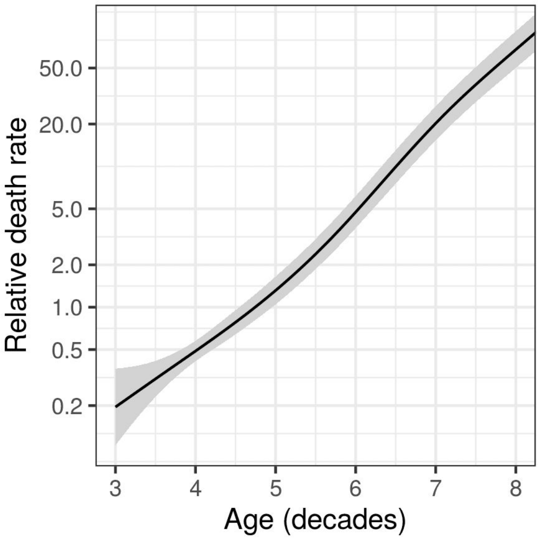


*Figure S7 - A restricted spline with 4 knots used to model the relationship between age as a continuous term and hazard of mortality in S-gene negative cases in an otherwise unconstrained model shows no obvious non-linearity.*

### Proportional hazards assumption

As described in the main text, we found that the assumption of constant hazard over time was violated; Figure S8 and Table S3. This is corroborated when we estimated the probability that the assumption was violated using the methods of Grambsch et al [1] and a further 3 methods for matched control studies, as presented in Xue et al [2]

**Table S3. Probability that the assumption of constant hazard over time was valid.**

| method | P(assumption valid) |
|---|---|
| cox.zph | 0.00459 |
| Event time correlation | 0.00452 |
| KM Estimate correlation | 0.00499 |
| Rank event time correlation | 0.00439 |

The proportional hazard assumption violation can be seen in the Kaplan Meier curve presented in the main paper ( and copied here for clarity). As the survival probabilities of S-gene positive and negative patients diverge around 14 days from the first positive test, we investigated whether we could resolve this violation of assumptions by fitting a model stratified by the time from first positive test result as a categorical variable of 0-14 days and 15-28 days.

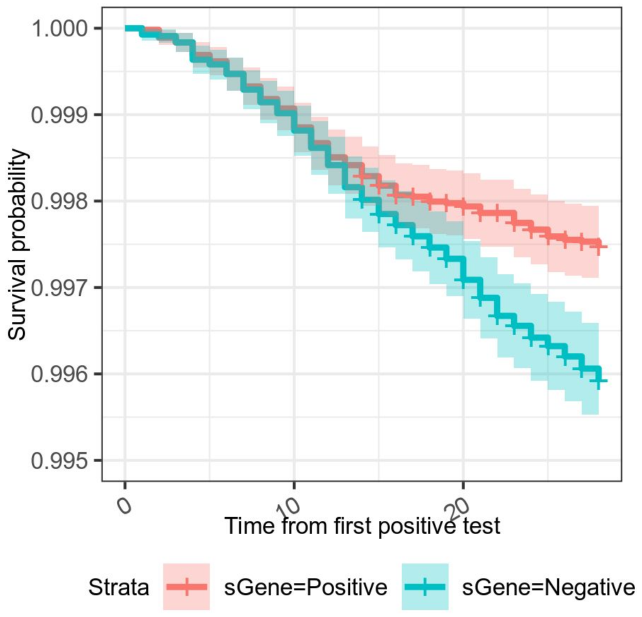


*Figure S8 - A Kaplan Meier curve of mortality of S-gene positive and S-gene negative COVID-19 infections. There is a divergence noted at 14 days.*

The hazard ratios for this model is presented in Table S4. These can be interpreted as saying that during the course of a COVID-19 infection, once the patient has survived past 2 weeks there is a relative improvement in their chance of surviving the next 2 weeks (HR 0.5). However this improvement in survival is seen principally in S-gene positive cases and S-gene negative cases continue to be at high risk of death into weeks 3 and 4 of their clinical course.

**Table S4. Cox proportional hazard model with time dependent terms, considering the impact of S-gene status on an early period (days 0-14) and a late period (days 15-28).**

| Model | Predictor | Value | Hazard ratio (95% CI) | p value |
|---|---|---|---|---|
| Early vs Late | Period | 0-14 (ref) | — | — |
| | | 15-28 | 0.51 (0.35 – 0.73) | <0.001 |
| | S-gene status | Positive (ref) | — | — |
| | | Negative (Period 0-14) | 1.23 (0.92 – 1.64) | 0.079 |
| | | Negative (Period 15-28) | 2.40 (1.66 – 3.47) | <0.001 |

For the time dependent model the proportional hazard assumption is not violated as demonstrated in table S5. Thus considering the follow up as 2 separate periods is enough to describe the increase in hazard ratio of mortality.

**Table S5. Probability that the assumption of constant hazard over time was valid with introduction of day 0-14 and day 15-28 periods.**

| method | P(assumption valid) |
|---|---|
| cox.zph | 0.756 |
| Event time correlation | 0.759 |
| KM Estimate correlation | 0.751 |
| Rank event time correlation | 0.758 |

Given this evidence that the hazard ratio is dependent on time, it is possible that the central estimate presented in the main paper will change as more data become available, and analysis which includes longer follow up becomes possible [3] depending on how the hazard ratio behaves past 28 days. This is in evolution and needs further investigation

The time dependence of mortality hazard has potential clinical significance. If S-gene negative COVID-19 infection is shown to be associated with later deaths it may be possible to modify this risk by earlier treatment or enhanced monitoring. This finding is something that can be validated in an unpaired retrospective observational cohort analysis, which would be powered to detect any subgroups for whom this may be a particular concern, and is a subject for future work.

### Supplementary material references


1 Grambsch PM, Therneau TM. Proportional Hazards Tests and Diagnostics Based on Weighted Residuals. *Biometrika* 1994;**81**:515–26. doi:10.2307/2337123

2 Xue X, Xie X, Gunter M, *et al.* Testing the proportional hazards assumption in case-cohort analysis. *BMC Med Res Methodol* 2013;**13**:88. doi:10.1186/1471-2288-13-88

3 Hernán MA. The Hazards of Hazard Ratios. *Epidemiol Camb Mass* 2010;**21**:13–5. doi:10.1097/EDE.0b013e3181c1ea43
